# A qualitative exploration into the role of illness perceptions in endometriosis-related quality of life

**DOI:** 10.1101/2023.03.16.23287347

**Authors:** Chloe Moore, Nicola Cogan, Lynn Williams

## Abstract

**Objectives:** Endometriosis is linked to adverse quality of life (QoL) outcomes. In the absence of effective treatment for endometriosis-related symptoms, supporting the QoL of those experiencing endometriosis is crucial. Illness perception (IP) interventions have prompted modest increases in QoL in several chronic conditions, yet IPs have not been comprehensively studied in relation to endometriosis. It is, therefore, necessary to examine the IPs held by individuals experiencing endometriosis to establish whether IP-based interventions might be useful in supporting QoL in this population. This research aims to gain an understanding of the IPs held by people experiencing endometriosis and their impact on QoL.

**Design:** Qualitative using one-to-one online semi-structured interviews.

**Methods:** Thirty individuals with endometriosis participated. Interviews sought to gain an understanding of participants’ experiences and perceptions in relation to living with endometriosis. Reflexive thematic analysis was used to develop themes.

**Results:** Three major themes were developed: (1) a life disrupted; (2) lost and fragmented sense of self; and (3) complex emotional responses. Largely negative IPs were held by individuals living with the condition which, along with endometriosis-specific symptoms and reduced functioning, fuelled fears for the future and reduced QoL.

**Conclusions:** Endometriosis-specific symptoms fuelled adverse QoL outcomes directly, and indirectly through moulding IPs. The disruption to the life trajectory associated with experiencing the condition as well as perceptions of control had a major impact on participants’ wellbeing, self-concept, and the varied emotional responses associated with experiencing endometriosis. IP-based interventions may support the wellbeing of those experiencing endometriosis whilst effective treatment is sought.

## Introduction

Endometriosis is a progressive, incurable condition characterised by the presence of endometrium-like tissue outside the uterus (Chapron et al., 2019). Commonly reported symptoms include chronic pelvic pain, menorrhagia, dyspareunia, sub-fertility and fatigue, although symptoms vary significantly between those diagnosed with the condition (Maddern et al., 2020; Ramin-Wright et al., 2018). Endometriosis affects approximately 1 in 10 women and people assigned female at birth globally (World Health Organization, 2021).

Endometriosis has a detrimental impact upon mental health and wellbeing (Vannuccini et al., 2017; Wang et al., 2021). Up to 64.4% and 63.5% of individuals diagnosed with the condition experience depression and anxiety respectively (Sepulcri & Amaral, 2009), whilst approximately 56% meet the clinical parameters for psychiatric diagnosis (Pope et al., 2015). Endometriosis is further associated with an adverse impact on quality of life (QoL; Della-Corte et al., 2020; Kalfas et al, 2022; Marinho et al., 2018), although there is ongoing debate surrounding the mechanisms by which endometriosis leads to reduced QoL.

Pain is the most prominent driver of endometriosis-related QoL outcomes (Broc et al, 2023; Facchin et al., 2015). As pain severity increases, as too does the likelihood of stress, anxiety and depression (Bullo & Hearn, 2021; Facchin et al., 2017), and reduced QoL (Culley et al., 2013). However, currently there is a lack of effective treatment for endometriosis-related pain and symptomology, so the contribution of other factors to QoL has been examined to ascertain how best to support individuals diagnosed with the condition. Reduced functioning (especially in work, sexual and social relationships; Nnoaham et al., 2011); diagnostic delays (Culley et al., 2013); and coping styles (González-Echevarría et al., 2019) have all been implicated in contributing to reduced QoL in endometriosis. Therefore, rather than one single factor underlying endometriosis-related QoL outcomes, QoL is likely determined by a complex interplay of several physical, social and psychological factors.

Qualitative research suggests that the way in which people perceive their illness, for example, perceived control surrounding the progression and impact of endometriosis and the anticipated consequences of experiencing the condition, is linked to QoL (Jones et al., 2004). A useful framework for comprehensively examining illness perceptions (IPs) is Leventhal’s (1997) common sense model of self-regulation (CSM-SR). In this model, IPs are conceptualised as a person’s beliefs and expectations relating to a health threat or medical condition. According to the CSM-SR, IPs work together with an emotional response to the health threat to drive behavioural responses and coping behaviours (Leventhal et al., 2016). This model situates IPs in 5 areas: 1) illness identity; 2) expected timeline of the health condition/threat; 3) the anticipated consequences of the health condition/threat; 4) the perceived cause of the condition; 5) perceived control and effectiveness of treatment in regulating or lessening symptoms (Leventhal et al., 2016). Moss-Morris et al. (2002) extended this framework to include a further 3 IP dimensions: 6) the extent to which an individual understands their condition; and 7) the emotional response to the health threat/condition; 8) concern surrounding the condition. Control was split into two distinct IPs: treatment control and personal control.

IPs are important drivers of mental health and QoL in several chronic conditions including fibromyalgia (van Wilgen et al., 2008), irritable bowel disease (Rochelle & Fidler, 2013) and rheumatoid arthritis (Hyphantis et al., 2013). Zhang et al. (2016) reported that, for individuals experiencing Crohn’s Disease, IPs directly influenced anxiety, depression and QoL. Specifically, perceiving negative consequences associated with Crohn’s, and framing the condition as uncontrollable increased the likelihood of depression, anxiety, and lowered QoL. Furthermore, interventions directly targeting IPs have led to modest improvements in treatment outcomes, mental wellbeing, and QoL in several conditions including diabetes, breast cancer and myocardial infarction (Alyami et al., 2021; Fischer et al., 2012; Sararoudi et al., 2016), suggesting that such interventions may support the QoL and wellbeing of individuals experiencing endometriosis.

IPs have not yet been comprehensively studied in relation to endometriosis. Previous qualitative literature, however, suggests that beliefs surrounding control and the consequences of endometriosis are related to QoL (Moradi et al., 2014; Young et al., 2015), with more negative perceptions increasing QoL detriments. This suggests that interventions focussed on reframing the IPs of individuals experiencing endometriosis may be beneficial in supporting QoL in the absence of effective treatment for endometriosis. However, endometriosis is dynamic and progressive with no known cure, and as such the emotions and cognitions associated with endometriosis are likely to evolve over the course of the condition.

Therefore, this study aimed to gain an understanding of the ways in which endometriosis is perceived and experienced by people diagnosed with the condition, and how these cognitions affect QoL. Findings will be compared against pre-defined IP categories to assess whether the illness-related beliefs of people experiencing endometriosis conform to or transcend these categories.

## Method

### Participants

Thirty participants were recruited from a pool of individuals who had completed a related survey as part of a broader project investigating factors associated with QoL in individuals with endometriosis. These participants were recruited from endometriosis support organisations and social media. A sampling matrix prioritising the recruitment of individuals with a range of ethnic backgrounds, ages, employment status’, household incomes, and educational attainment was used to ensure a diverse sample.

Participants were aged between 20 and 55 years (M=35.6, SD=9.49). Additional participant demographics are displayed in table 1. Individuals were eligible to participate if they had been formally diagnosed with endometriosis (e.g., through laparoscopic investigation), were over the age of 18 and resided in the UK or Ireland. Participants had experienced endometriosis symptoms for 4 – 40 years (M=14.83 years, SD=9.18), and had been diagnosed for an average of 5 years (SD=6.97). Further information on the nature of participants’ endometriosis is presented in table 2. Participants were given pseudonyms to preserve their anonymity.

**Table 1.** Participant demographics

|  | N | % |
| --- | --- | --- |
| **Ethnicity** |  |  |
| White British | 20 | 66.7% |
| Indian | 2 | 6.7% |
| African | 2 | 6.7% |
| Another Mixed Background | 2 | 6.7% |
| Pakistani | 1 | 3.3% |
| Asian and White | 1 | 3.3% |
| Another Ethnic Background | 1 | 3.3% |
| Another White Background | 1 | 3.3% |
| **Relationship Status** |  |  |
| Married | 10 | 33.3% |
| Cohabiting with partner | 10 | 33.3% |
| Single | 9 | 30% |
| Widowed | 1 | 3.3% |
| **Educational attainment** |  |  |
| Undergraduate / Bachelors degree | 10 | 33.3% |
| Postgraduate degree | 6 | 20% |
| Secondary education to GSCE/O-levels/National 5 or equivalent | 4 | 13.3% |
| Secondary education to Highers/A-level or equivalent | 3 | 10% |
| Diploma of Higher Education/Foundation Degree/Higher National Diploma/NVQ level 5/level 5 diploma or equivalent | 3 | 10% |
| Left school with no qualifications | 2 | 6.7% |
| Completed secondary school to National 3/4 or standard grade | 1 | 3.3% |
| Prefer not to say | 1 | 3.3% |
| **Employment status** |  |  |
| Employed full-time | 11 | 36.7% |
| Disabled or unable to work | 6 | 20% |
| Employed part-time | 5 | 16.7% |
| Self-employed | 3 | 10% |
| Unemployed, looking for work | 2 | 6.7% |
| Employed on a zero hours or casual contract | 1 | 3.3% |
| Full-time student | 1 | 3.3% |
| Part time student | 1 | 3.3% |
| **Country of residence** |  |  |
| England | 17 | 56.7% |
| Scotland | 12 | 40% |
| Wales | 1 | 3.3% |

**Table 2.** Participant endometriosis information

|  | N | % |
| --- | --- | --- |
| **Treatment** |  |  |
| NHS | 16 | 53.3% |
| Some private, some NHS | 13 | 43.3% |
| Completely private | 1 | 3.3% |
| **Co-morbid condition** |  |  |
| Yes | 18 | 60% |
| No | 12 | 40% |
| **Surgery** |  |  |
| Had surgery | 24 | 80% |
| Not had surgery | 6 | 20% |
| **Number of surgeries** |  |  |
| 1 | 10 | 33.3% |
| 2 | 8 | 26.7% |
| 3 | 3 | 10% |
| 4 | 2 | 6.7% |
| 5 | 1 | 3.3% |
| **Trying for baby** |  |  |
| Yes | 12 | 40% |
| No | 17 | 56.7% |
| Prefer not to say | 1 | 3.3% |

### Sample size

A sample size of 30 was deemed appropriate to provide an in-depth, reflective account of participants’ experiences (Farrugia, 2019). Data saturation is not considered due to the incompatibility of this concept with reflexive thematic analysis (RTA), which was adopted for this study (Braun & Clarke, 2019; 2021). In RTA, codes and themes are fluid and organic, and subsequently there is no clear point at which codes and themes cease to materialise as posited by the concept of data saturation.

### Data collection

Upon receiving ethical approval from the host institution, the sampling matrix was used to identify potential participants who had previously indicated their interest in participating in an interview. An information sheet was sent to selected individuals via email and those interested provided written consent to be interviewed. Interviews were semi-structured and facilitated by the first researcher online. Interviews were audio recorded and lasted 42 to 90 minutes (M=62 minutes). Participants reiterated their consent verbally before the commencement of the interview. Reflexive notes were taken throughout. Following the interview, participants were debriefed and offered a £20 Amazon e-voucher as compensation for their time.

A topic guide consisting of open-ended questions and prompts was developed. This included broad questions relating to participant’s beliefs about their condition and more specific questions informed by the pre-existing IP model. The topic guide was piloted with 2 participants to ensure that questions were relevant and comprehensive. During the interviews, participants were asked to describe the impact of endometriosis on their lives, before answering questions surrounding their perceptions of endometriosis. Topics included the consequences of living with endometriosis, the emotional impact associated with the condition, and perceived control over endometriosis.

### Analysis

Data was analysed in line with Braun and Clarke’s (2006; 2019) guidelines for RTA, due to the theoretical flexibility associated with this approach, and its capacity to reduce large quantities of data into comprehensive, accessible themes that provide a coherent, nuanced account of participant experiences (Braun & Clarke, 2012). An inductive approach was first adopted to develop themes out-with a theoretical framework, before a deductive approach was taken to compare the identified themes to pre-established IP dimensions (Leventhal et al., 1997). Throughout the analytical process, a reflexive journal was kept by the first researcher to note thoughts, feelings, and assumptions relevant to this process. Initially, the first author read through each transcript several times whilst noting prominent ideas deriving from participant accounts. NVivo was used to organise the data into 156 codes. Similar and duplicate codes were merged, and others were redefined leaving 124 codes. Potential themes were derived by grouping related codes together. Themes were reviewed based on their relevance to the research question before they were defined and named. The final stage of analysis involved discussion amongst all authors regarding the appropriateness of the themes and their definitions in relation to the research question, before three themes were selected and finalised for this report.

## Results

Three major themes were constructed: (1) a life disrupted; (2) lost and fragmented sense of self; and (3) complex emotional responses. Each theme mapped onto multiple pre-existing IP dimensions (figure 1).

**Figure 1.**
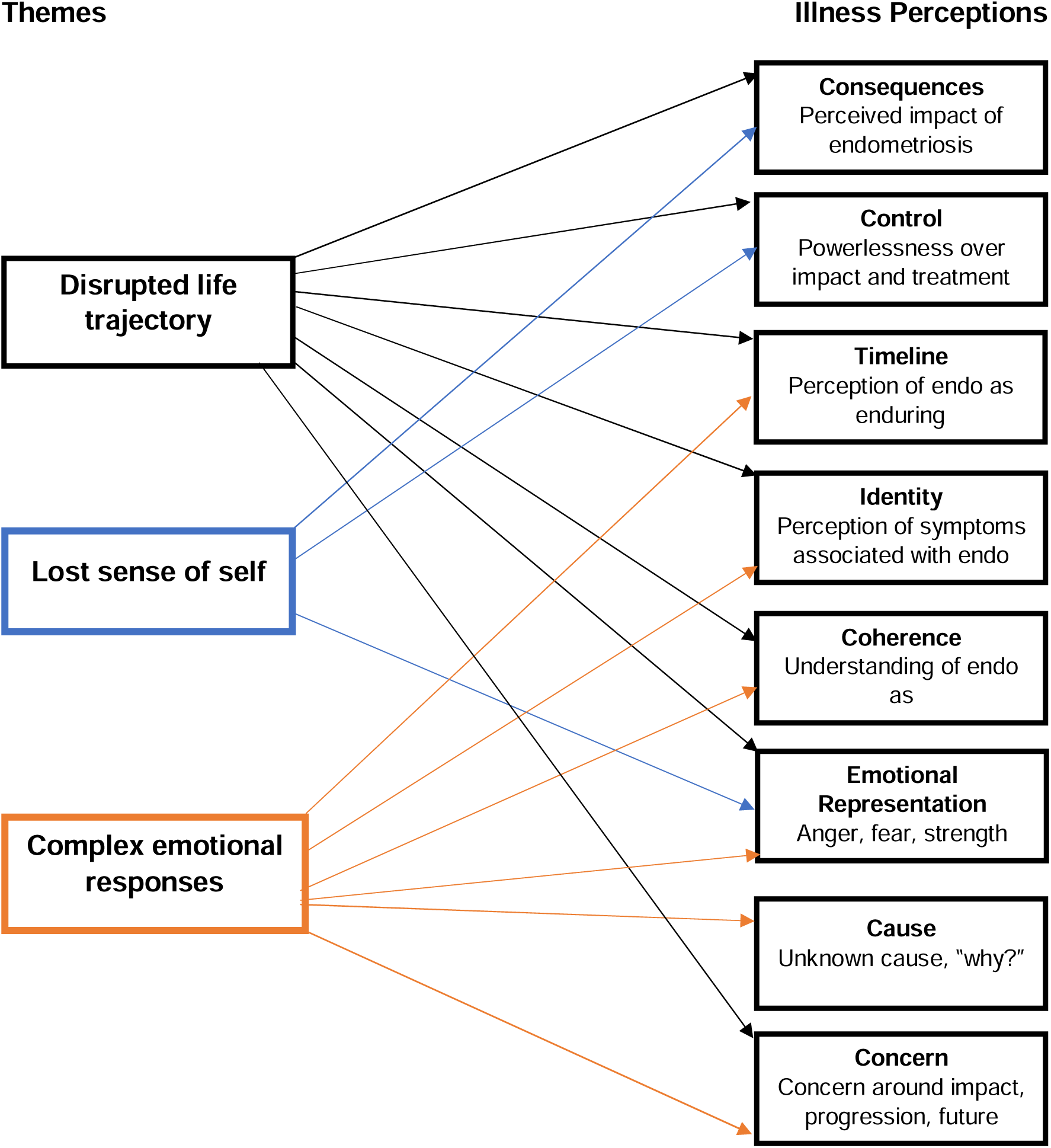
Relationship between identified themes and illness perceptions

### Theme 1: A life disrupted

Participants all referred to the multiple and varied life disruptions that were an inevitable consequence of living with endometriosis. Many disclosed that endometriosis prevented them from living the life they wanted and felt that their potential in life had not been realised due to the debilitating symptoms they experienced. What constituted participants’ potential varied between accounts. Often, potential was defined in terms of career or education goals, although some described their potential in relation to their relationships or fertility. There was a sense of lost time and missed opportunities attributed to endometriosis:

> “*I feel like it’s [endometriosis] kinda taken away my youth and it’s taking away from all of the things that I aspire to do and that I could have done and could have achieved*.” [Ash].

This sense of loss was woven through participant accounts, encompassing several life domains including education, work, relationships and, often, day-to-day functioning. Participants stressed that no aspect of their lives was left untouched by the impact of endometriosis:

> “*It’s an everyday thing that impacts on my actual ability to just function in life*.” [Morgan].

The language used by participants implied a sense of powerlessness attributed to endometriosis, which manifested itself in participants’ life trajectories. Several perceived endometriosis as “controlling” [Alina] their lives, and viewed their life trajectories as dictated by the progression of the condition:

> “*I felt like all the choices I would want to make about my own life, that I should have made, it felt like it was taken away by endometriosis. […] I feel like I’m not in control of my life, this illness is*.” [Billie].

Perceived disruption to life trajectories often prompted a negative emotional response. Participants described feelings of sadness and frustration related to their circumstances, with some experiencing symptoms of anxiety and depression because of endometriosis-related life disruption(s). Adverse impacts on QoL and mental wellbeing were emphasised by several participants, largely facilitated by functioning detriments triggered by endometriosis (e.g., social functioning, work functioning, day-to-day functioning):

> “*It’s actually not the pain that’s the worst part of the disease, it’s the impact that it does have on my life and how it prevents me from doing things that has affected my mental health more than anything.*” [Emily].

This impact on wellbeing was associated with several life domains including work and relationships. Negative mental health outcomes were particularly pertinent for participants experiencing infertility or uncertainty around their fertility. It must be noted that participants differed widely in their thoughts, feelings and experiences with regards to fertility and parenthood, with several participants childfree by choice, some actively trying to conceive and others who had successfully conceived. Nonetheless, several participants described feelings of helplessness, powerlessness, and anxiety related to their experiences with fertility, particularly when conceiving was difficult or unsuccessful:

> “*I worry will I ever have children, what lies ahead for me?*” [Indra]

Participants often voiced a sense of uncertainty and fear surrounding the future, and this sentiment was not exclusive to fertility. There was a general underlying apprehension regarding the future woven through participant accounts. This apprehension was often fuelled by historic disruptions to participants’ life trajectories across several life domains (e.g., work, relationships) as well as the incurable nature of endometriosis, leading many to speculate that their symptoms would last forever. Participants displayed great awareness of the progressive nature of their condition, often driving fears that endometriosis-related symptoms might worsen over time and dictate their future outcomes:

> “*It’s just this idea of, it can grow as it likes, it has no cause that’s known, there’s no treatment plan, the pain is excruciating and it will never go away. And I think that to me felt like, this thing is going to kind of colonise my life*.” [Polly]

However, participant responses were complex and diverse, and correspondingly some did not share the same apprehensions surrounding the future as described above. Several described “taking one day at a time” [Jackie] and attempting to focus on the present rather than the future [Skye] to prevent fears surrounding the future from taking hold:

> “*I don’t look too far ahead because if I look at this time 6 months ago I was in a completely different place so, you know, I just take every day as it comes and make the most of it*.” [Violet]

Similarly, some participants had learned to find ways to live with the life disruptions associated with endometriosis, for example by entering jobs with flexible working patterns, or keeping diaries of their symptoms and triggers to pre-empt endometriosis flare-ups and prepare, as far as possible, for impending symptoms. After disclosing the disruption caused by endometriosis to their sexual functioning, one participant described how re-framing their definition of sex had minimised the disruption on their relationship:

> “*We haven’t had penetration in our sex life for the full 2 years of our relationship and it hasn’t impacted on our love for each other and our intimacy because he doesn’t want to see me in pain and I’m at a stage where I’m confident enough within myself that I’m not willing to put myself through pain for anyone else either so we work with ways together. Em, we can both experience pleasure that doesn’t involve me being in agony and crying because that isn’t fun for anyone. Re-framing that definition [of sex] has helped me to feel better about myself as a woman.*” [Emily]

As demonstrated by this extract, restructuring pervasive and negative perceptions around the destructive nature of endometriosis led to improvements in self-esteem for this participant, which in itself is linked to overall wellbeing and QoL (Martinsen et al., 2021).

Clear through each participant’s account was their resolve to regain some of the control over their life trajectory that was thought lost through endometriosis:

> “*I’m going to be able to live with this, it’s not going to take my life.*” [Sarah]

### Theme 2: Lost and fragmented sense of self

Participants generally felt that their identity was moulded and driven by endometriosis. Several participants described their sense of self as “lost” [Violet], often stating that they felt like a “different person” [Robin] owing to the impact of endometriosis on their lives:

> “*I don’t think I’ll ever be the person I was before, I think this [endometriosis] has changed me forever*.” [Becky].

Some participants described the heavy emotional burden associated with experiencing a progressive, often debilitating condition. There was a sense that endometriosis slowly eroded participant’s sense of self, that the longer this emotional burden was carried, the more significant the impact on their self-concept:

> “*You feel like you’re not yourself anymore. You’re someone, you’re someone different. You sort of become the illness in a way.*” [Billie].

This sense of the self as lost to endometriosis was often interlinked with feelings of vulnerability elicited by the symptoms associated with endometriosis. For many participants, pain and fatigue progressed with the condition, provoking a shift in self-perception. Some defined themselves as increasingly “sick” [Indra], “unwell” [Alex], and there was a sense amongst some that endometriosis had stripped away previously salient aspects of their identity:

> “*I did change and, um, I became a victim. Um, and that wasn’t me before, you know, I was always independent, stood on my own two feet and I didn’t rely on anybody and with, with the pain and everything else I became a different person*.” [Robin]

Perceptions of the self as increasingly “unwell” [Alex], along with functioning detriments engendered by endometriosis symptoms led to shifts in specific aspects of the identity. Femininity and sexual identity were particularly impacted by endometriosis in this participant group and were intrinsically linked, in that specific symptoms (e.g., pain during sex, fatigue) eroded participant’s sexual drive which dismantled perceptions of femininity. This sense of diminished femininity then impacted sexual drive and desire:

> “*I just physically can’t [have sex]. I don’t feel feminine, I don’t feel sexy because I’m in pain and living in oversized jogging bottoms*.” [Nathalia]

Several participants perceived themselves as “less of a woman” [Billie; Emily] due to the impact of endometriosis-related symptoms on their sexual functioning and feminine identity. The sense that femininity was lost to endometriosis had a marked impact on the self-esteem of several participants:

> “*I’m really bloated so that means I can’t wear that nice dress that I want to wear, I can’t wear heels. But now I have to go and find something that doesn’t dig into my stomach and is more floaty and, like, in terms of like, self-confidence as a woman, that’s taken a big hit.*” [Nathalia]

However, not everyone who experiences endometriosis identifies as feminine or female. One non-binary participant described the impact of living with endometriosis on their own self-perception. Living with symptoms such as menstrual bleeding and chronic pelvic pain activated a sense of gender dysphoria in them, and left them feeling “confused”, “isolated” and with a fragmented sense of their own gender identity:

> “*[diagnosis] didn’t help with the old gender identity because they’re very much, this is a woman’s disease, this is a woman’s illness. This is a thing that happens to women, people with uteruses. And I’m just over here like, oh no.*” [Morgan]

Changes to the self-concept engendered by endometriosis were not only linked to symptomology, but the broader treatment of endometriosis in both societal and medical settings. There was a general sense that symptoms were minimised and dismissed by others, particularly healthcare professionals. Several participants described the internalisation of these minimised symptoms, that what they were experiencing was characteristic of regular menstruation and the result of a “low pain threshold” [Iona; Robin]. This led many participants to question their knowledge and expertise in their own bodies. Many accepted that the symptoms they were experiencing were ‘normal’ and felt instead that it was their response to these symptoms that was abnormal:

> “*I was just thinking, I’m imagining these pains, there is no pain here, I’m just imagining it.*” [Charlie]

There was often a sense of an internal struggle to control the self-concept amongst participants, in which internalised notions that their symptoms were not real or were a ‘normal’ part of menstruating were pitted against an internal sense that their symptoms were real and valid:

> “*You feel like something’s there but you keep getting told that nothing’s there and then it’s this anxiety of, I’m imagining things. I don’t know what’s real and what’s not anymore*.” [Indra]

Despite the wide-reaching impact of endometriosis on the sense of self, there was a general determination amongst participants not to allow endometriosis to seize their identity completely:

> “*I think, well it doesn’t define me, this is just something that I deal with and I cope with*.” [Sarah]

Additionally, some participants reflected on the positive ways in which endometriosis had shaped their identity, specifically highlighting patience, resilience and strength:

> “*it’s only recently that I’ve looked back on everything and thought you know what, I am strong, I can see my identity as more, you know, I do identify as someone who does, I’ll push for things and I’m brave and I’ll talk about things and I’m not shy about it and, um, you know, I think that’s sort of changed my identity*.” [Alina]

### Theme 3: Complex emotional responses

Participants’ endometriosis-related experiences prompted several emotions. Emotions varied widely but can be separated into two categories: i) endometriosis as an emotional burden; ii) endometriosis as a facilitator of emotional strength. Participants generally described emotions in each of these categories, highlighting the complexity and sometimes conflicting feelings surrounding endometriosis.

### 3.1 Endometriosis as an emotional burden

Frustration was the most prominent emotion described in relation to participants’ experiences of endometriosis. This often stemmed from knowledge of the incurable and progressive nature of endometriosis, and the lack of effective treatment available. The unknown cause of endometriosis also gave rise to feelings of frustration:

> “*How are there people that don’t end up suffering? Obviously you wouldn’t wish it on anyone but it’s just that understanding of why certain people get it and why other people don’t and it makes you feel frustrated that it does end up being you that experiences it*.” [Jenny]

Widespread misunderstanding and the minimisation of participant experiences in medical settings was also a common source of frustration:

> “*It’s been frustrating that no-one would take me seriously, frustrating that lead times on appointments were too long, frustrating, frustrating that no-one seems to understand it, frustrating that, you know, it’s something we’ve known about for hundreds of years and yet we still don’t know anything about it.*” [Becky]

Frustration led to feelings of anger for many participants. Anger was often intertwined with the perceived negative impact of endometriosis on the life trajectory and identity, linked with feelings of powerlessness:

> “*I am raging inside that I’ve got to be kind of forced into a position of being weak and not being able to do what I want to do*.” [Reece]

Feelings of sadness were also described by several participants. As above, sadness tended to revolve around a sense of powerlessness and lack of control over the condition. Participants sometimes voiced a sense of being “attacked” [Ava] by their own body which led to sadness and hopelessness:

> “*Sometimes I’m like I can’t believe my body is betraying me, it’s like really just rubbish, why does, you know, so that’s quite, I would say, a little bit upsetting* [crying].” [Evelyn]

Guilt was another prominent emotion throughout participant accounts. Guilt tended to surround the impact of endometriosis on relationships, for example, being physically unable to engage in sexual activity with intimate partners or rejecting social invitations from friends due to endometriosis-related symptoms. Participants with daughters also often shared a sense of guilt and dread at the prospect of their children inheriting the condition:

> “*What kills me is I’ve just had a baby, and when I found out it was a girl it was definitely in my head that this is something that I’m now going to pass on to her and she’s now going to have to live with this and that part made me upset*.” [Sarah]

Furthermore, participants often described feelings of loneliness, often prompted by a sense that their experiences were misunderstood by large swathes of society due to a lack of understanding and education surrounding endometriosis:

> “*I have went for the better part of about 13 years going no-one else experiences what I experience. You know, and having no one else that understands it is very, very isolating*.” [Mira]

For some participants, the emotional pain they experienced due to endometriosis progressed into longer-term mental health concerns. Many described enduring long periods of low mood and symptoms of depression:

> “*It [endometriosis] affected my mental health big time. I woke up in the morning and just felt like there was this black cloud above my head and I didn’t want to get up, I just wanted to hide away.*” [Billie]

### 3.2 Emotional strength stemming from endometriosis

Contrarily, some participants described finding emotional strength through their experiences of living with endometriosis. This strength was often forged through establishing coping mechanisms to mitigate the mental health impact of endometriosis:

> “*I think it’s had to make me a stronger person because I’ve just had to deal with it, it’s just something that I’ve, that’s part of my life*.” [Sarah]

The notion of “dealing with” endometriosis implies a sense of control over the condition, indicating that emotional strength may be derived from challenging the feelings of powerlessness that are so often linked to endometriosis.

Participants often derived feelings of empowerment and strength through using their experiences to support and advocate for others. For several participants, this gave value and purpose to their experiences:

> “*I’ve done a lot of work like helping other people which has gave me a bit of like purpose and again something good that’s come out of it where at least I’ve been able to help other people whether it’s to give advice and support or just to listen and tell them ‘I understand what you’re going through’*.” [Emily].
>
> “*It’s almost like having a little piece of wisdom that you get from unfortunate circumstances.*” [Alina]

One participant described endometriosis as giving them a sense of “pride” [Casey]. Benefit finding was common amongst participants, indicating a determination to mitigate against the negative emotional impact associated with the condition.

## Discussion

This study is the first to qualitatively explore endometriosis-related IPs and their relation to QoL amongst individuals experiencing endometriosis. An inductive and deductive approach to analysis allowed for IPs to be considered both organically and within a theoretical framework.

Broadly, the findings reflect previous research suggesting that endometriosis has a detrimental impact on the QoL and wellbeing of those experiencing the condition (e.g., Wang et al., 2021). There were, however, disparities within participant accounts regarding the extent to which endometriosis impacted aspects of QoL, with some recounting a pervasive, debilitating effect on their lives, and others describing a more manageable, fluctuating impact. This nuance in participant experience is likely associated with disparities in endometriosis symptomology. Although the findings of the current study conform to the notion that endometriosis symptomology is inherently and irreversibly linked to QoL, they also add to the existing literature by highlighting additional mechanisms by which endometriosis may impact QoL and wellbeing, specifically by moulding and shaping IPs which, within this participant group, were linked to dimensions of QoL such as life trajectory, the self-concept, and emotional impact. IPs were shaped both directly by endometriosis symptomology and indirectly through functioning detriments. Figure 2 provides a visual representation of this relationship as derived from participant accounts within this study.

**Figure 2.**
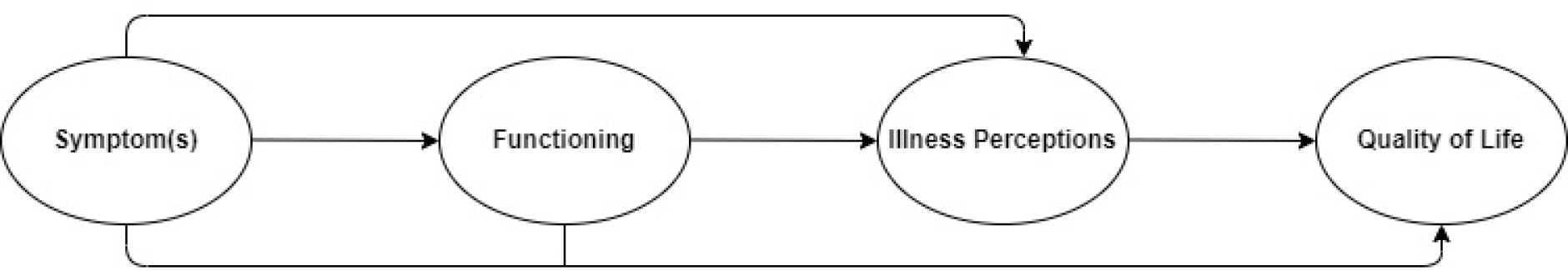
Relationship between endometriosis symptoms, functioning, IPs and QoL

Each of the inductively identified themes mapped on to multiple pre-defined IPs (figure 1). Previous research has already demonstrated the pervasive, negative impact of endometriosis on life domains such as relationships, sleep, work and education, and sex and fertility (Halici et al, 2023; Missmer et al., 2021). Theme 1 encapsulates these effects, demonstrating the wide-ranging negative perceived and actual consequences of endometriosis on participants’ life trajectories. Participants often highlighted specific symptoms such as pain and fatigue as the cause of disruption to their expected life trajectories, indicating a lack of control over endometriosis and the subsequent impact on their lives. This is perhaps unsurprising, given the incurability of endometriosis and research demonstrating that treatment is ineffective in the long-term for many individuals (Nirgianakis et al., 2020). Indeed, participants demonstrated a strong awareness and knowledge of their condition, including the incurable nature of endometriosis and potential progression of their symptoms, and this coherence was instrumental in cultivating feelings of powerlessness in relation to endometriosis. Furthermore, there was a general sense that endometriosis symptoms would persist throughout the lifespan. Perceptions of the enduring timeline of endometriosis were linked to fears associated with the consequences of endometriosis on the life trajectory as well as perceptions of control over the condition. Research suggests that endometriosis symptoms persist even after menopause (Secosan et al., 2020), potentially fuelling the fears for the future voiced by several participants. The perceived consequences of endometriosis on the life trajectory also prompted a strong emotional response from many participants, who described detrimental wellbeing effects stemming from the disruption and anticipated disruption to their lives, including anxiety and sadness. However, importantly, some participants described re-framing their perceptions around the consequences of endometriosis, leading to improvements in their wellbeing and QoL. It is important that future research investigates this potential link further to establish whether interventions to address IPs may be beneficial for individuals experiencing endometriosis.

Corresponding with previous qualitative research suggesting a link between endometriosis and the identity (Cole et al., 2021), theme 2 highlights a fragmented and lost sense of self attributable to participants’ experiences of endometriosis. Although identity is a pre-defined illness perception, ‘identity’ within this theme transcends the definition offered by the CSM-SR, in which it is centred around perceptions of the symptoms associated with the condition rather than the sense of self (Leventhal et al., 2016). In the current study, identity refers to the broader self-concept and theme 2 explores participants’ perceptions of the way in which this is moulded, driven, and changed by endometriosis. This theme is interlinked with theme 1, as many of the perceived changes to identity stemmed from the impact of endometriosis on specific life domains such as work and relationships. This is in-keeping with research demonstrating that the sense of self is intrinsically linked to social aspects including career choice (Fryers, 2006), familial relationships (Anderson & Chen, 2002) and sex (Hensel et al., 2011). In turn, the self-concept is linked to QoL in several chronic conditions (e.g., Octari et al., 2020), suggesting that IPs may indirectly impact endometriosis-related QoL through shaping the identity. Additional research is required to examine this potential link further. Within this study, participants used terms such as “lost” to describe their identity, implying a sense of powerlessness and loss of control surrounding their sense of self. However, using the term “lost” rather than “broken” or “gone” suggests a sense that the self-concept may be recovered, as found among people living with other chronic conditions (Cogan et al, 2016; Golub et al, 2014; Vann-Ward et al, 2017). This finding implies an underlying hope that control of the identity might be regained from endometriosis. This corresponds with the dichotomy observed within some participant accounts, in which the sense of self was described as driven by endometriosis but, simultaneously, there was a determination to not allow endometriosis to take over the identity.

Examining participant’s perceptions of their identity through an IP lens revealed shared experiences amongst participants such as the internalised trivialisation of endometriosis-related symptomology, which corresponds to broader social themes including the treatment of women’s health conditions in medical and societal settings. There is vast sociological discourse on the treatment of women’s health conditions that corresponds with participant’s accounts of the minimisation of their symptoms at both societal and medical levels (e.g., Alexander et al., 2020). Within the current study, participants often questioned their knowledge and expertise in their own bodies, with some doubting their experiences and even their ‘sanity’. In this, many participants appeared to experience a sense of externalised self-perception (Jack & Dill, 1992) in which they viewed themselves through the lens of others. This is described as an act of self-silencing and has been linked to endometriosis in previous qualitative research (Cole et al., 2021).

A strong emotional response to endometriosis was woven throughout participant accounts, and this is described in theme 3. Feelings of anger and frustration correspond to previous qualitative research where they are often intertwined with endometriosis-specific factors such as treatment effectiveness and diagnostic delay (Jones et al., 2004). Within this study, the emotional response was often interlinked with perceptions of control, coherence, consequences and the anticipated longevity of endometriosis symptoms. Negative emotional responses were most prominent throughout participant accounts, corresponding with previous literature suggesting that frustration, fear and sadness are common amongst people experiencing endometriosis (Young et al., 2015). However, perhaps surprisingly, some participants described positive emotions associated with endometriosis, emphasising resilience, pride and strength cultivated by endometriosis. This was often linked with a sense of hope for the future and participants finding value in their experiences by supporting, educating, and advocating for others. In this participant sample, benefit finding was a commonly used strategy to lessen the emotional impact of endometriosis. The impact of benefit finding on emotions and wellbeing has not yet been researched in endometriosis and therefore constitutes an important area for future research.

IPs in this participant sample could be matched to each of the pre-defined IPs as described in the CSM-SR (figure 1). Most prominent in this sample were perceptions of control and consequences, which were clearly linked to participants’ life trajectories and sense of self. Less clear however is the role of the illness identity (i.e., how endometriosis is viewed by those living with the condition) or the perceived cause of endometriosis. As there is no known cause for endometriosis, perceptions around causation may not be particularly strong within this population which may be reflected in the results of this study. However, recent research suggests that many individuals experiencing endometriosis hold subjective views of the cause of their condition (Münch et al., 2022), and in the current research the lack of a known cause itself did prompt an emotional response in some participants. Therefore, future research could endeavour to establish whether there is a link between perceptions of endometriosis cause and QoL and/or wellbeing.

Considering the findings of this research, namely that the experiences of participants are linked to IPs, and that shifts in some IPs appear to prompt positive QoL and psychological outcomes, it is possible that IP-based interventions may partially mitigate the detrimental impact of endometriosis on QoL outcomes. This is not to say that psychological intervention can replace effective treatment, but that it may support the wellbeing of individuals diagnosed with endometriosis whilst reliable treatment is sought. Due to the dearth of research on this topic, future research could assess IPs with a large sample of individuals using pre-established measures of IPs such as the revised illness perception questionnaire (Moss-Morris et al., 2002) to investigate further the appropriateness of studying endometriosis within a CSM-SR framework before corresponding interventions are trialled.

## Strengths and limitations

To our knowledge, this is the first study to qualitatively consider the IPs of individuals living with endometriosis. This paper extends current knowledge on the mechanisms underlying adverse QoL outcomes in people experiencing endometriosis by suggesting that IPs may contribute to QoL alongside stronger predictors of wellbeing such as pain. However, this study must be viewed in light of its limitations as well as its strengths. Firstly, participants were largely recruited through social media channels and support groups, indicating that many had actively sought support for their condition. People involved in support groups may hold views unreflective of the wider population in two distinct ways: i) they may have worsened symptomology and more negative experiences leading them to seek support; ii) they may have a more positive outlook regarding their endometriosis diagnosis due to increased support and coping mechanisms.

Additionally, the interview topic guide was underpinned by the CSM-SR and many questions related to pre-existing IPs. Therefore, although an inductive approach was taken in constructing themes, the information yielded from the interviews may have been heavily slanted towards the CSM-SR’s depiction of IPs. Therefore, important IPs held by participants but existing out-with this theoretical framework may have been missed and the role of pre-established IPs over-emphasised. However, in investigating IPs within a pre-established framework, this research lays the groundwork for future investigation into the role of IPs in endometriosis-related outcomes by suggesting that these cognitions likely contribute to QoL and psychological wellbeing outcomes.

## Conclusion

This study highlights the complex and dynamic nature of the IPs held by individuals experiencing endometriosis. Endometriosis-specific symptoms such as pain were the main driver of QoL detriments within this population, and these symptoms and their associated impact cultivated and moulded endometriosis-related IPs. Whilst effective treatment continues to be sought for endometriosis-related symptoms, it is important that research continues to investigate the factors that may mitigate the detrimental impact of endometriosis on QoL and wellbeing. These findings offer clear indications that interventions based on endometriosis-related IPs may support the QoL of individuals experiencing endometriosis, and suggests that future research explore the link between IPs and QoL in endometriosis further.

## Data Availability

Research data are not shared due to privacy and ethical concerns.

## Data availability statement

The data that support the findings of this study are not available due to privacy and ethical restrictions.

## Acknowledgements

This project is funded by a Scottish Graduate School for Social Science Economic and Social Research Council Studentship award (Project Reference: ES/P000681/1). Special thanks to the endometriosis support groups who assisted with the design and recruitment of this research.

